# Detecting Medication Mentions in Social Media Data Using Large Language Models

**DOI:** 10.1101/2025.05.16.25327791

**Authors:** Guillermo Lopez-Garcia, Dongfang Xu, Graciela Gonzalez-Hernandez

## Abstract

The automatic extraction of medication mentions from social media data is critical for pharmacovigilance and public health monitoring. In this study, we present an end-to-end generative approach based on instruction-tuned large language models (LLMs) for medication mention extraction from Twitter. Reformulating the task as a text-to-text generation problem, our models achieve state-of-the-art results on both fine-grained span extraction and coarse-grained tweet-level classification, surpassing traditional sequence labeling baselines and previous best-performing systems. We demonstrate that fine-tuning Flan-T5 models enables efficient and accurate extraction while simplifying the architecture by eliminating complex multi-stage pipelines. Additionally, we show that lexicon-based filtering further improves performance by reducing false positives. Our models are publicly available, providing high-performing and efficient tools for large-scale pharmacological analysis of social media data.

## 1. Introduction

Social media posts are valuable sources of patient-generated data, offering real-time, large-scale insights into public health [1]. These posts support novel approaches to syndromic surveillance [2], pharmacovigilance [3], and monitoring medication abuse [4]. An essential step in using social meidia data for pharmacoepidemiology is the automatic detection of medications from them, which enables large-scale analysis of how drugs are discussed, perceived, and used in real-world settings [5].

Over the last decade, techniques for medication detection have been advancing, thanks in part to recent shared tasks including automatic detection of posts mentioning a drug name from SMM4H-2018 Task 1 [6] and SMM4H-2020 Task 1 [7], and automatic extraction of medication mentions from tweets from BioCreative-VII Task 3 [8]. The SMM4H shared tasks are framed as binary text classification challenges. While these tasks do not directly extract medication mentions, they serve as a crucial filtering step to reduce the volume of irrelevant posts, which is essential because a large proportion of drug name mentions appear in advertisements or posts by bots, and many drug names are ambiguous. BioCreative-VII Task 3 is a named entity recognition (NER) task, where the objective is to identify the exact text span corresponding to each medication mention.

Most participants in the SMM4H-2018 and 2020 shared tasks used neural network classifiers with hierarchical tweet representations [6], as well as BERT-based models as classifiers [9]. The current state-of-the-art (SOTA) system for both SMM4H tasks is the top-performing system submitted to SMM4H-2020 [10], which is based on an ensemble of 20 BERT-based classifiers [11]. For the BioCreative shared task, most participants fine-tuned BERT-style language models, such as Megatron-BERT-345M [12] or PubMedBERT [13, 14] to perform token-level classification. While the core approach was similar, participants varied in their use of data augmentation strategies, ensembling techniques, and text preprocessing methods. The SOTA system in the BioCreative shared task employed a two-stage architecture: a tweet-level classification module using an ensemble of seven fine-tuned BERTweet-large models [15], followed by a span-level extraction component based on Splinter—a BERT-based model optimized for span-level question answering [16]—to identify the exact medication mentions.

Despite their strong performances, systems with pipeline modules or ensemble techniques pose significant efficiency challenges for large-scale text mining in pharmacovigilance and digital epidemiology, where rapid processing of hundreds of thousands of tweets is essential. For the pipeline-based approach, one additional drawback is the risk of cascading errors, where misclassifications at the tweet-level stage can negatively affect downstream span-level extraction.

To address the limitations of prior work, we propose an end-to-end approach that leverages large language models (LLMs) to automatically extract medication mentions from tweets. Unlike prior pipeline-based methods that separate classification and extraction into distinct stages, our approach reformulates the task as a single-step text generation problem. Specifically, we fine-tune Flan-T5 models [17], a lightweight family of LLMs, to perform direct extraction of medication mentions from tweet text. Our method achieves SOTA performance on the BioCreative VII Task 3, while offering substantial improvements in both simplicity and computational efficiency. Additionally, we evaluate our proposed approach on SMM4H-2018 and SMM4H-2020 datasets for binary tweet classification. Despite not being explicitly trained for text classification, our generative approach outperforms existing task-specific models, achieving new SOTA results on both benchmarks. We make our models publicly available at https://github.com/guilopgar/Medication-Detection-LLM, providing the community with high-performing and efficient systems for large-scale pharmacological analysis of social media data.

## 2. Methods

We developed an end-to-end system for extracting medication mentions using instruction fine-tuning of Flan-T5 models [17]. Flan-T5 extends the original T5 (Text-to-Text Transfer Transformer) architecture [18] by incorporating instruction tuning, which enables the model to follow a wide range of natural language commands. This makes Flan-T5 particularly effective for prompt-based, generative approaches to information extraction.

To leverage this capability, we framed medication extraction as a question-answering (QA) task, following the instruction tuning format used in Flan-T5. As shown in Table 1, each prompt begins with a task-specific instruction that defines the expected output format. The model is then given a tweet and asked to identify any drug names, medications, or dietary supplements mentioned. During training, the model receives both the prompt and the corresponding answer—a structured list of medication mentions—allowing for supervised fine-tuning. At inference, the model generates this list directly from the prompt.

**Table 1.** Prompt template used for medication mention extraction in our generative approach, which frames the task as a QA problem. Each prompt includes an instruction, the tweet content, a question, and the expected model output.

|  |  |
| --- | --- |
| **Instruction** | You are given a tweet followed by a specific question asking about the content of the tweet. Your objective is to identify and list any drug names, medications, or dietary supplements mentioned in the tweet. If one or more are mentioned, list each distinctly, separated by a comma. If none are mentioned, return an empty list [].<br>Input: Tweet: <tweet content><br>**Question: What are the drugs, medications or dietary supplements mentioned in the tweet?**<br>Output: <model output> |
| **Example 1** | Input: Tweet: Bedtime snack, benadryl, New Girl, and acetaminophen. The party is getting real.<br>Question: What are the drugs, medications or dietary supplements mentioned in the tweet?<br>Output: [benadryl, acetaminophen] |
| **Example 2** | Input: Tweet: @LWR I miss the front desk. Plus I don't work on Monday's or Tuesday's. U all should call me. I'll work the desk<br>Question: What are the drugs, medications or dietary supplements mentioned in the tweet?<br>Output: [] |

### 2.1. Lexicon-based Filter

To improve precision, we applied a lexicon-based postprocessing filter from the Kusuri system [5], which removes extracted mentions not found in a predefined drug lexicon. Each extracted mention was compared to the lexicon using case-insensitive partial matching. A mention was retained only if at least one of its tokens matched a token from any drug entry in the lexicon. We used the same lexicon as in [19], which includes known drug names and their common variants from RxNorm.

## 3. Datasets

Table 2 summarizes the data statistics of the three corpora used in our study. For each corpus, we followed the standard train/test splits and task setups as defined in their respective shared tasks.

**Table 2.** Number of tweets in the training, validation, and test sets for each annotated corpus. For each set, we also include the percentage of tweets containing at least one drug (shown in round brackets).

| Corpus | Task | Train | Validation | Test |
| --- | --- | --- | --- | --- |
| BioCreative VII Task 3 [8] | Medication extraction | 88,988 (0.2%) | 38,137 (0.2%) | 54,482 (0.2%) |
| #SMM4H 2018 Task 1 [6] | Tweet classification | 9,622 (52%) | – | 5,382 (53%) |
| #SMM4H 2020 Task 1 [7] | Tweet classification | 69,272 (0.3%) | – | 29,687 (0.3%) |

The BioCreative VII Task 3 corpus includes 181,607 tweets with gold-standard (GS) annotations marking the exact character offsets of drug names, medications, and dietary supplements. It reflects the naturally imbalanced distribution of drug mentions on Twitter, with only 0.2% of tweets containing at least one medication mention [5].

The SMM4H-2018 corpus consists of 15,004 tweets, originally annotated for binary classification to indicate whether a tweet contains one or more drug mentions. Approximately 50% of the tweets are positive, resulting in an artificially balanced class distribution. Although initially designed for classification, the BioCreative VII Task 3 organizers later released a span-annotated version of the training set, enabling its use for medication mention extraction.

The SMM4H-2020 corpus contains 98,959 tweets, with only 0.3% mentioning at least one medication. Like BioCreative VII, this dataset reflects the natural, highly imbalanced distribution of drug mentions on Twitter [5].

### 3.1. Evaluation

We evaluated model performance on the medication mention extraction task using the official metrics from BioCreative VII Task 3. Specifically, we report Precision (P), Recall (R), and F1-score (F1) under two evaluation settings: 1) strict evaluation, where a prediction is considered correct only if it exactly matches GS text span; and 2) overlapping evaluation, where a prediction is correct if it overlaps with any portion of a GS span.

For the SMM4H-2018 and 2020 tweet classification tasks, we also follow the official metrics and report P, R, and F1 scores for the positive class, defined as tweets that mention one or more medications. We compared our proposed approach against established baselines and state-of-the-art (SOTA) systems for each dataset:

#### Token Classifiers

We implemented token classification models using two BERT-based architectures:

**RoBERTa-large** and **BERTweet-large**, both trained with the standard BIO tagging scheme [20].

#### BioCreative-Baseline

A hybrid system combining a lexicon-based classifier with a BERT-base token classifier for medication mention extraction [19].

#### BioCreative-SOTA

A two-stage pipeline consisting of a BERTweet-large ensemble for tweet classification, followed by a BERT-based QA model for medication extraction [21].

#### SMM4H-2018-Baseline

The top-performing system from SMM4H-2018 Task 1, which used neural classifiers with hierarchical tweet representations [6].

#### SMM4H-2018-SOTA

A binary tweet classifier based on the BERT-base model [19].

#### SMM4H-2020-Baseline

The Kusuri system [5], a hybrid model combining deep neural networks with lexicon-based filtering, served as the official baseline for SMM4H-2020 Task 1 [7].

#### SMM4H-2020-SOTA

An ensemble of 20 BERT-based binary classifiers, which achieved the best performance in the SMM4H-2020 shared task [10].

We evaluated two instruction-tuned generative models—**Flan-T5-large** and **Flan-T5-XL**—selected for their balance between instruction-following ability and computational efficiency. For medication extraction, we fine-tuned both Flan-T5 models and token classifiers on a combined dataset comprising the updated SMM4H-2018 Task 1 training set and the BioCreative VII Task 3 corpus. We applied lexicon-based postprocessing to outputs from both generative and token classification models. For tweet classification, we directly evaluated the above Flan-T5 and token classification models without additional fine-tuning on tweet classification data.

We tuned the hyperparameters based on models’ performances on the BioCreative VII Task 3 validation set. Both Flan-T5 models were trained with a learning rate of 5 × 10^™5^, a batch size of 32, and 10 training epochs. We conducted all experiments on a single NVIDIA A100 GPU with 80 GB of VRAM.

### 3.2. Postprocessing

Since our generative model outputs a comma-separated list of medication mentions (as shown in Table 1), we implemented a postprocessing step to map each predicted mention back to its character span in the original tweet. We used a case-insensitive exact string matching procedure: each predicted mention was lowercased and searched within the lowercased tweet text. All matching substrings were assigned start and end character offsets. Mentions that could not be matched to any part of the tweet were discarded. This mapping allowed us to compute character-level, span-based evaluation metrics.

To derive tweet-level predictions from the generative models and baseline systems trained on the extraction task, we used a simple rule: if models found at least one medication mention for a tweet, it was labeled as positive; otherwise, it was labeled as negative.

## 4. Results

### 4.1 Medication mention extraction

Table 3 presents the performance of all evaluated models on the medication mention extraction task using the BioCreative VII Task 3 test set.

**Table 3.**
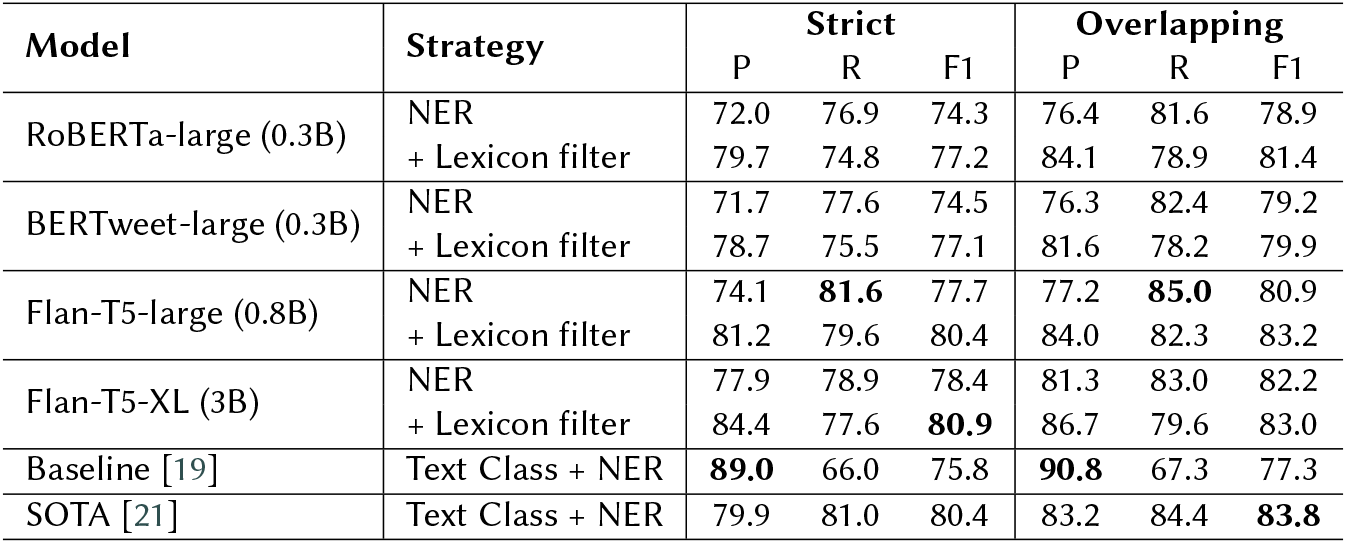
Performance of medication mention extraction models on the test set from the BioCreative VII Task 3 (Strict and Overlapping evaluation). For each metric, the highest value is in bold (P: Precision, R: Recall, F1: F1-score).

The Flan-T5 models consistently outperformed both token classifiers based on RoBERTa-large and BERTweet-large. Under the strict evaluation setting, the Flan-T5-XL model achieved the highest F1-score of 80.9 when combined with lexicon-based filtering. According to the F1-score under the strict evaluation setting, which was considered as the official evaluation metric of the task, Flan-T5-XL with lexicon filtering surpassed the performance of the SOTA system reported in the shared task (80.4).

While the baseline models exhibited strong recall, their precision was lower compared to the generative models. The application of lexicon-based filtering consistently improved precision across all models, often leading to notable gains in overall F1-score. For instance, RoBERTa-large improved from 74.3 to 77.2 F1-score under strict evaluation after lexicon filtering.

### 4.2. Tweet Classification

Table 4 summarizes the results of the binary tweet classification task on the SMM4H-2020 and SMM4H-2018 test sets. Despite not being explicitly trained for tweet-level classification, the generative Flan-T5 models outperformed the performance of dedicated classification models.

**Table 4.**
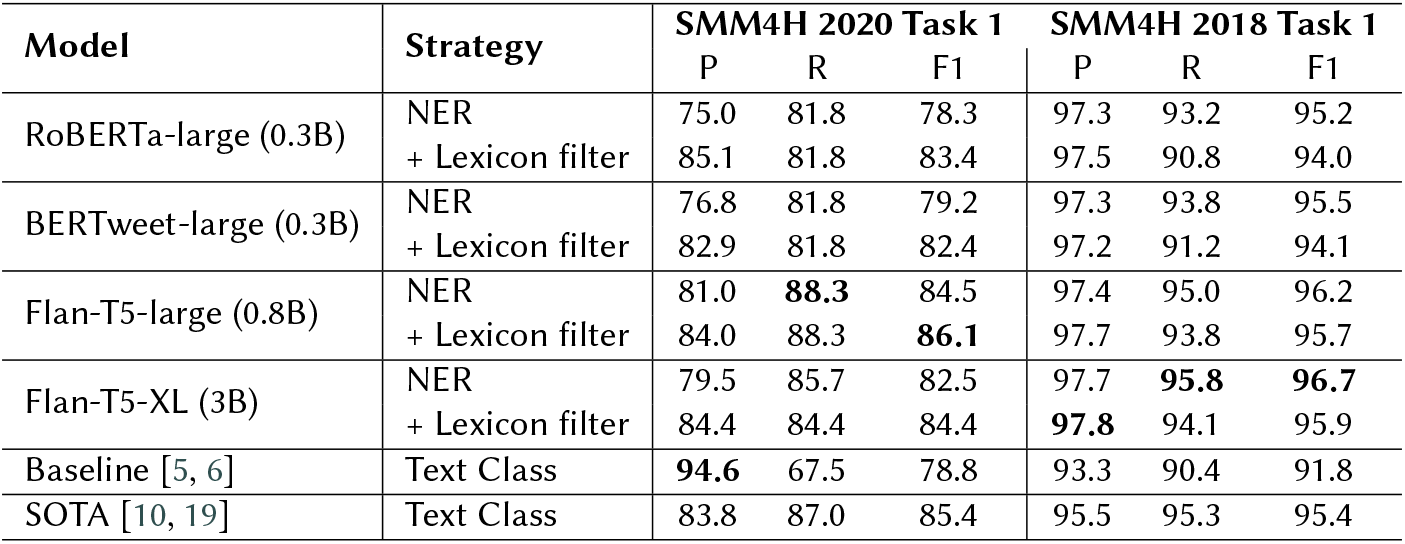
Performance of medication mention classification models on #SMM4H 2020 and #SMM4H 2018 test sets. For each metric, the highest value is in bold (P: Precision, R: Recall, F1: F1-score).

On the highly imbalanced SMM4H-2020 test set, Flan-T5-large combined with lexicon filtering achieved the highest F1-score of 86.1, outperforming the traditional sequence labeling baselines and the SOTA system reported for this task. On the more balanced SMM4H-2018 test set, Flan-T5-XL achieved the highest F1-score of 96.7, also surpassing all baseline systems and the SOTA.

The application of lexicon filtering proved beneficial for improving precision without substantial losses in recall on the SMM4H-2020 test set. However, for the SMM4H-2018 Task 1 dataset, where models already achieved higher precision than recall, lexicon filtering did not provide additional benefit and slightly reduced F1-scores for all models.

## 5. Discussion

We also investigated the performance of Flan-T5 models under a few-shot prompting strategy [22], where no task-specific fine-tuning was performed. In this setup, four illustrative examples were dynamically incorporated into the prompt for each input tweet. These examples were selected based on their semantic similarity to the target tweet, computed using cosine similarity over sentence embeddings generated by the `all-mpnet-base-v2` model on Sentence Transformer [23].

**Table 5.** Performance of Flan-T5 models with few-shot prompting and fine-tuning on the test set from the BioCreative VII Task 3.

| Model | Strategy | Strict |  |  | Overlapping |  |  |
| --- | --- | --- | --- | --- | --- | --- | --- |
|  |  | P | R | F1 | P | R | F1 |
| Flan-T5-large (0.8B) | Few-shot prompting | 44.2 | 51.7 | 47.6 | 48.0 | 56.5 | 51.9 |
|  | Fine-tuning | 74.1 | **81.6** | 77.7 | 77.2 | **85.0** | 80.9 |
| Flan-T5-XL (3B) | Few-shot prompting | 68.8 | 43.5 | 53.3 | 71.0 | 44.6 | 54.8 |
|  | Fine-tuning | **77.9** | 78.9 | **78.4** | **81.3** | 83.0 | **82.2** |

Despite the careful selection of relevant examples, the few-shot prompting approach consistently underperformed compared to fully fine-tuned models. Specifically, while the Flan-T5-XL model achieved an F1-score of 78.4 under strict evaluation after fine-tuning, its few-shot counterpart only reached 53.3. Similar trends were observed for the Flan-T5-large model, where the fine-tuned model obtained an F1-score of 77.7 under strict evaluation, and its few-shot counterpart only achieved 47.6. These results highlight the limitations of relying solely on in-context learning for complex information extraction tasks, particularly in low-resource and highly imbalanced domains such as pharmacovigilance on social media. Fine-tuning remains critical for adapting language models to the linguistic challenges posed by noisy, informal text and domain-specific vocabularies.

### 5.1. Limitations and future work

Although our proposed generative approach achieved SOTA results in medication mention extraction and tweet-level classification, several limitations should be acknowledged. First, the extreme class imbalance inherent in the medication mention extraction task from social media data poses significant challenges for both model training and evaluation. While instruction fine-tuning led to strong performance, our current approach did not incorporate dedicated data augmentation techniques to mitigate this imbalance. Future work will explore the integration of data augmentation strategies, such as the synthetic generation of medication-mentioning tweets and controlled paraphrasing of existing examples, to enrich the positive class and improve model robustness.

Additionally, our study focused exclusively on models from the Flan-T5 family due to their strong instruction-following capabilities and computational efficiency, critical factors for large-scale analysis of social media data. In future work, we will investigate the use of more advanced LLMs [24] to address the medication mention extraction task. Specifically, we aim to explore their potential to overcome the limitations observed with few-shot learning when using Flan-T5 models.

## 6. Conclusion

In this study, we developed an end-to-end generative approach based on instruction-tuned LLMs for the automatic extraction of medication mentions from tweets. Our models achieved SOTA performance on both fine-grained span extraction and coarse-grained tweet-level classification tasks, surpassing traditional sequence labeling baselines and prior SOTA systems. Additionally, our end-to-end generative approach benefits from a simplified and more efficient architecture, eliminating the need for complex multi-stage pipelines.

## Data Availability

All data used in this study are publicly available through the organizers of the #SMM4H 2018, #SMM4H 2020, and BioCreative VII Shared Tasks

https://github.com/guilopgar/Medication-Detection-LLM

